# Diagnostics and prognostic potential of current biomarkers in heart failure with preserved ejection fraction: a systematic review and meta-analysis

**DOI:** 10.1101/2020.04.18.20070482

**Authors:** Hao Chen, Michael Chhor, Benjamin Rayner, Kristine McGrath, Lana McClements

## Abstract

**Aims:** Circulating biomarkers are commonly used in diagnosis and prognosis of heart failure with preserved ejection fraction (HFpEF) in clinical practice. However, the diagnostic and prognostic potential of current biomarkers in HFpEF remain unclear.

**Methods and results:** We conducted a search of the PubMed, Web of Science, MEDLINE and SCOPUS (1900 to January 2020) databases of all diagnostic (n=1,104) and prognostic (n=53,497) biomarkers investigated in people with HFpEF. B-type natriuretic peptide (BNP) displayed satisfactory sensitivity (0.81, 95% CI: 0.76 to 0.85; I^2^=0) and specificity (0.86, 95% CI: 0.82 to 0.89; I^2^=16.9%) for the diagnosis of HFpEF. Natriuretic peptides (NPs), including N-terminal pro BNP (NT-proBNP) and BNP, were associated with over two-fold increased risk of mortality (NT-proBNP: HR=2.27, 95% CI: 1.69 to 3.06, I^2^=87.6%; BNP: HR=3.01, 95% CI: 1.27 to 7.21, I^2^=97.2%), hospitalisation (NT-proBNP: HR=3.54, 95% CI: 2.83 to 4.43, I^2^=83.4%), and a composite event of both (NT-proBNP: HR=2.55, 95% CI: 2.13 to 3.05, I^2^=78.1%; BNP: HR=2.28, 95% CI: 1.42 to 3.69, I^2^=75.8%) in people with HFpEF. Interestingly, Galectin-3 (Gal-3) (sensitivity: 0.70, 95% CI: 0.63 to 0.75, I^2^=86.7%; specificity: 0.78, 95% CI: 0.69 to 0.85, I^2^=68.6%) and soluble suppression of tumorigenicity 2 (sST2) (sensitivity: 0.58, 95% CI: 0.52 to 0.64, I^2^=88.1%; specificity: 0.59, 95% CI: 0.49 to 0.68, I^2^=69.5%) showed limited diagnostic potential of HFpEF.

**Conclusion:** Amongst currently available biomarkers, BNP remains the most reliable diagnostic marker of HFpEF. Although there was high heterogeneity between the studies included, BNP or NT-proBNP could also have promising prognostic potential in HFpEF.

## INTRODUCTION

Heart failure (HF) is becoming an increasingly prominent disease in developed countries, placing a burden on both patients and healthcare systems. HF affects 38 million people worldwide and in Australia alone there are currently 480,000 people diagnosed with this condition, with rising prevalence.^1^ HF is a complex syndrome characterised by abnormal cardiac structure and function that impairs the ability of heart to fill and eject blood at normal pressure. In line with this definition, current clinical guidelines classify HF into two subtypes based on left ventricular ejection fraction (LVEF) of the patients. An LVEF lower than 50% with or without clinical signs of HF is defined as HF with reduced ejection fraction (HFrEF), whereas LVEF from 50% and above in the presence of structural heart disease or diastolic dysfunction (DD) is defined as heart failure with preserved ejection fraction (HFpEF).^2^ HFpEF occurs in approximately 50% of patients with HF, and it is associated with similar mortality as HFrEF.^3^ HFpEF remains a challenging condition to manage as it is a phenotypically diverse syndrome, often difficult to diagnose. Whilst currently available treatments such as angiotensin-converting enzyme inhibitors (ACEIs), angiotensin II receptor blockers (ARBs), mineralocorticoid receptor antagonists (MRAs), diuretics, beta-blockers (BBs) are suitable for the management of patients with HFrEF, these are not effective for those with HFpEF.^4^

Circulating biomarkers are employed regularly in diagnosis and prognosis of HF and have additional roles in providing better understanding of the underlying pathophysiology of HFpEF, which could lead to the development of more effective therapies and improved patient outcomes. NPs, including N-terminal pro-b-type natriuretic peptide (NT-proBNP) and b-type natriuretic peptide (BNP), are currently recommended by clinical guidelines for diagnosis ^5, 6^ and prognosis ^6^ of HFpEF. In addition, Gal-3 and sST2 are emerging as reliable biomarkers for risk stratification of HFpEF.^6^ The use of current biomarkers is limited in determining the subtypes of HF and in predicting morbidity and mortality rates. Therefore, we conducted meta-analyses, which for the first time, comprehensively assesses the diagnostic and prognostic potential of the most commonly utilised biomarkers in the context of HFpEF.

## METHODS

### Data sources and searches

Two separate systematic searches were conducted to assess the diagnostic and prognostic accuracy of biomarkers in HFpEF using the following databases: PubMed, Web of Science, MEDLINE and SCOPUS (1900 to January 2020). Literature searches were performed using the following terms: “HFpEF and biomarker”, “HFpEF and biomarker and mortality”, and “HFpEF and biomarker and hospitalisation”. Full list of terms used is provided in the Supplementary material online (*Appendix S1*).

### Study inclusion criteria

The assessment of the biomarkers used in the diagnosis of HFpEF was carried out using the published data from observational studies assessing the diagnostic accuracy of individual biomarkers, which discriminate between cohorts with and without HFpEF. In terms of prognostic assessment of individual biomarkers, studies with primary outcomes defined as overall mortality, hospitalisation, and/or a composite event of both, in patients with HFpEF, were included. For the assessment of diagnostic biomarkers, studies were selected only if sensitivity and specificity of individual biomarker was reported, whereas for prognostic biomarkers, studies had to have reported hazard ratio (HR) associated with a biomarker in predicting the aforementioned primary outcomes. The HRs were required to be obtained based on dichotomous data comparisons between the two sides of biomarkers’ cut-off values. Studies in which HRs were obtained based on continuous data comparisons (e.g. HR per log increase in biomarker level) were excluded.

### Data extraction

Two independent investigators (H.C. and M.C.) extracted data from included studies. Disagreements were resolved by consensus with a third investigator (L.M.). The recommendations of PRISMA guidelines ^7^ and another relevant guideline ^8^ were followed for critical study reviews and data extraction.

For diagnostic biomarker assessment, true positive (TP), true negative (TN), false positive (FP) and false negative (FN) values of individual biomarkers to diagnose HFpEF, were extracted. For prognostic biomarker assessment, the HR values with 95% confidence intervals (CIs) associated with individual biomarker’s ability to predict the pre-specified outcomes, were extracted. The covariate adjustments of HRs were also recorded, if applicable.

### Quality assessment

The included studies were assessed for quality independently by three co-authors (M.C., B.R. and K.M.) using validated Quality Assessment for Diagnostic Accuracy Studies-2 (QUADAS-2) ^9^ and Quality in Prognosis Studies (QUIPS) ^10^ tools for diagnostic and prognostic biomarkers, respectively. Results were compared between assessors and, in case of disagreement, individual studies were discussed to achieve consensus. The quality scores based on QUADAS-2 and QUIPS tools for each study are listed in *Appendix S2*, Supplementary material online.

### Data analysis

The meta-analyses for diagnostic biomarkers was performed using sensitivity and specificity values of each biomarker, which discriminate between cohorts with and without HFpEF (controls vs. HFpEF). The estimated sensitivity and specificity of the diagnostic biomarkers were calculated using TP, TN, FP and FN values. The sensitivity and specificity of those biomarkers extracted from four or more independent studies, were pooled and analysed using Meta-DiSc 1.4 (Meta-DiSc, Madrid, Spain) and STATA 16.0 (STATA Corporation, College Station, Texas, USA) to generate forest plots and hierarchical summary of receiver operating characteristic (HSROC) curves, respectively. Forest plots were generated only for those diagnostic biomarkers that were investigated in at least three independent studies. For those diagnostic and prognostic markers extracted from less than three studies, we only recorded the sensitivity and specificity, and/or HRs estimates of the original studies without pooling the data.

We also analysed the prognostic accuracy of individual biomarkers by pooling HRs from a minimum of three independent studies. The univariable HRs were preferably selected, when not available, multivariable HRs with the least number of adjustments were used. The HRs with 95% CIs were estimated by random-effect model forest plots by STATA 16.0. Bias was assessed by the Egger’s regression test.

## RESULTS

### Search results

The search for HFpEF diagnostic biomarkers yielded 6,145 articles, of which 18 ^s1-s18^ were included; the search for HFpEF prognostic biomarkers yielded 2,440 articles, of which 26 ^s15, s19-s43^ met the inclusion criteria (*Figure 1*) (see Supplementary material online, *Appendix S3* for references). Most selected studies were designed either prospectively (n=20) ^s1-s3, s9-s13, s17- s20, s28, s37-s43^ or retrospectively (n=18) ^s4-s8, s14-s16, s25, s26, s29-s36^, whereas five ^s21-s24, s27^ were post hoc analyses of randomised controlled trials (RCTs). One study was included in both diagnostic and prognostic biomarker assessment^s15^. The vast majority of included studies utilised a 50% LVEF as a diagnostic criterium to define HFpEF whereas LVEF cut-off values for diagnosis of HFpEF were lower than 50% in eleven ^s1, s8, s20-s24, s28, s38, s42, s43^ of them. In total, 1,104 patients with HFpEF were included in diagnostic biomarkers assessments and 53,497 patients in prognostic biomarkers assessments. Patients were free of valvular diseases in all included studies. Most of the prognostic biomarker studies except four ^s25, s30, s40, s42^ reported baseline medications, with ranges across the studies as follows: ACEIs or ARBs (7.6%-88.1%), MRAs (15%-53%), BBs (17%-82.3%), digoxin (10%-88%), statins (27.4%-64%), diuretics (16%-100%).

**Figure 1.**
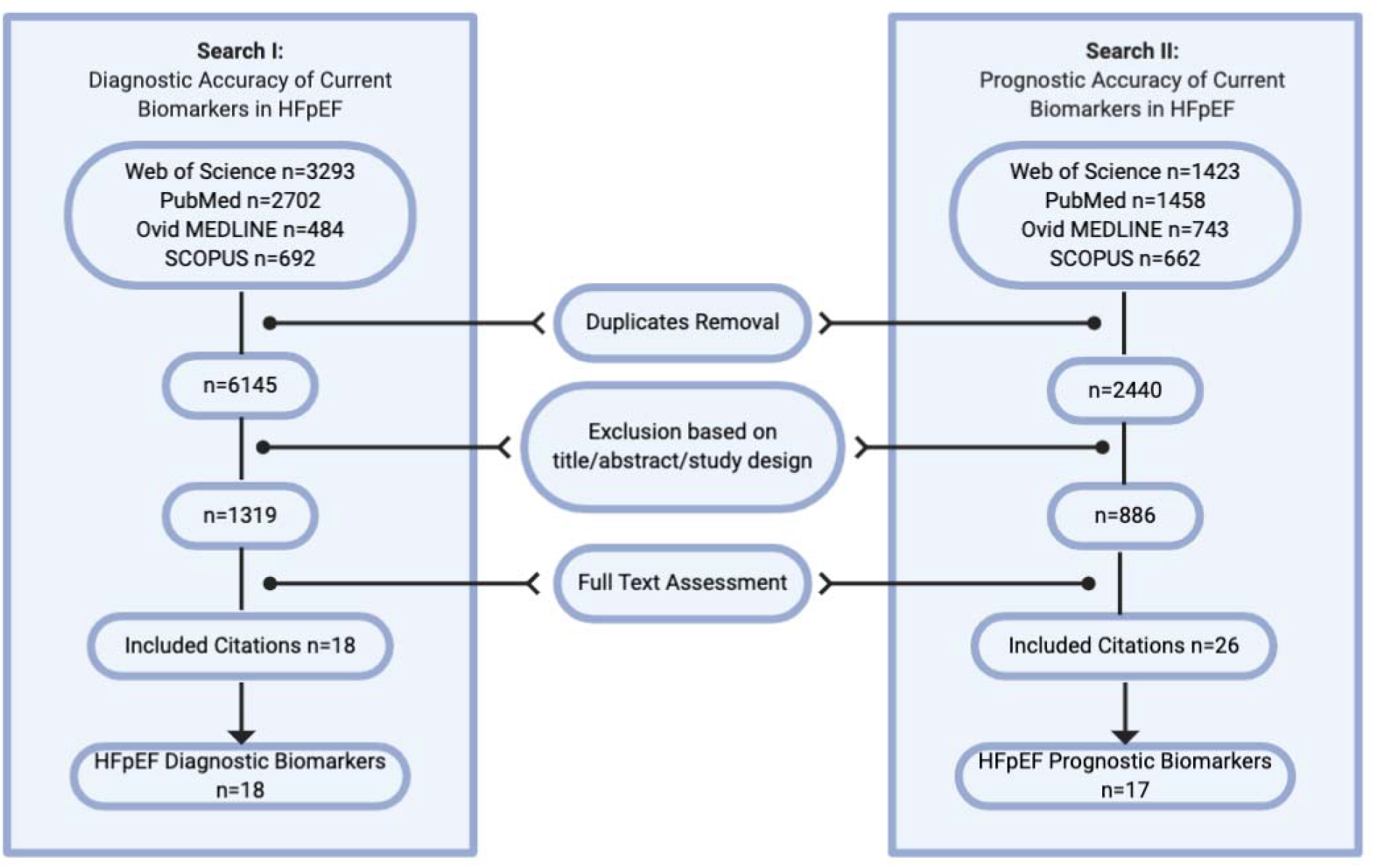
Preferred Reporting Items for Systematic Reviews and Meta-Analyses (PRISMA) guidelines flow diagram.

Overall, selected studies yielded a total of 18 different diagnostic markers and 17 different prognostic markers. Most of the studies evaluated NPs in both diagnosis (n=12) ^s1-s6, s8-s13^ and prognosis (n=15) ^s19-s26, s29-s35^ of HFpEF. The cut-off values of identical biomarkers were heterogeneous as were the follow-up periods.

### NT-proBNP as a biomarker of diagnosis and prognosis in HFpEF

In relation to the studies using NT-proBNP as a diagnostic biomarker of HFpEF (n=6) ^s1-s6^, three studies ^s1, s2, s5^ were conducted in Europe, and three studies ^s3, s4, s6^ were conducted in Asia. In the primary analysis where all six studies were included, NT-proBNP showed estimated pooled sensitivity of 0.69 (95% CI: 0.65 to 0.73) and specificity of 0.92 (95% CI: 0.89 to 0.94) in the diagnosis of HFpEF (*Figure 2A* and *2B*). However, there was significant heterogeneity in the sensitivity (*I*^2^=83.7%) and specificity (*I*^2^=83.5%) of NT-proBNP between the studies.

**Figure 2.**
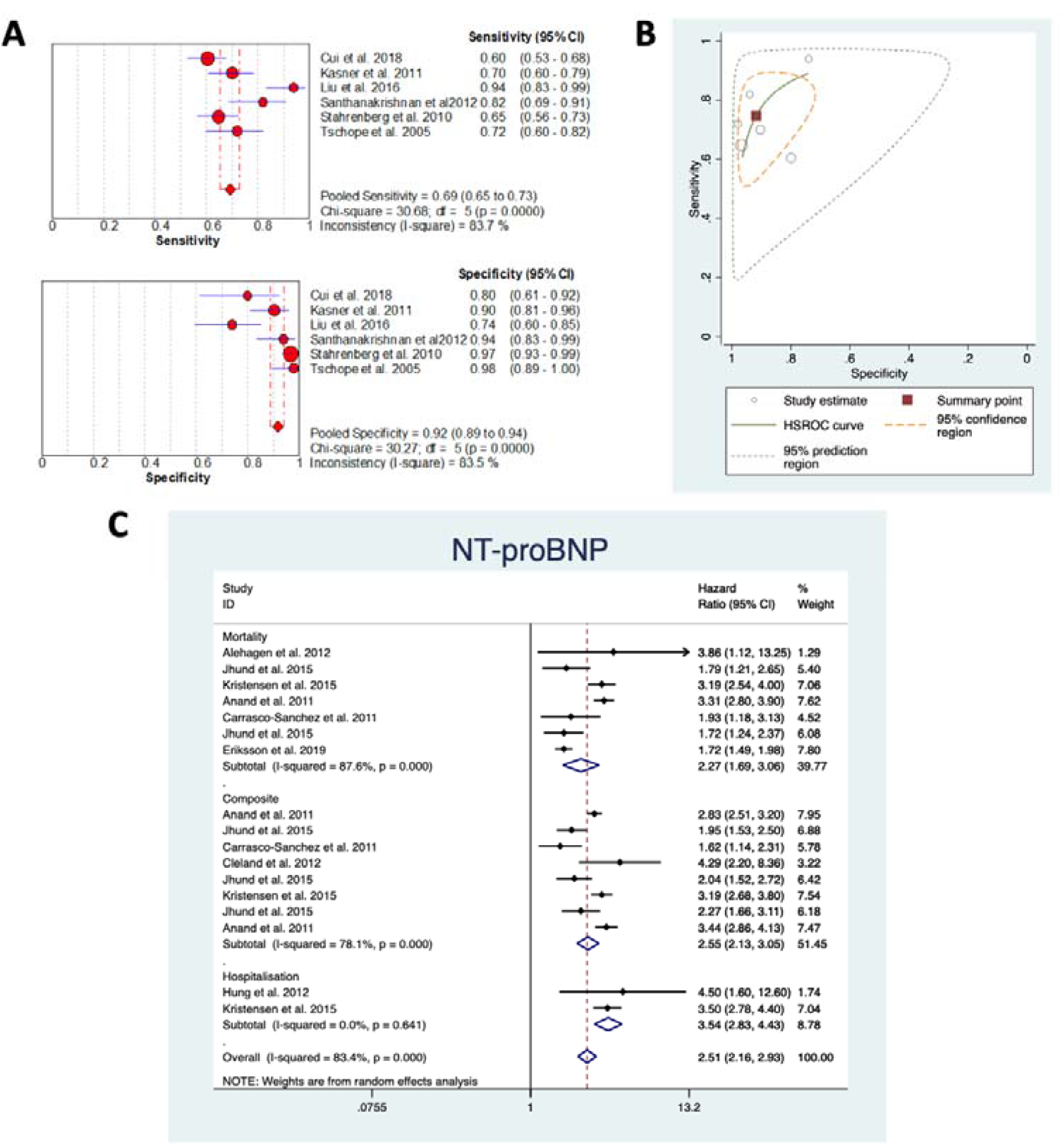
Sensitivity, specificity, and hazard ratio, of NT-proBNP extracted from all studies using NT-proBNP for diagnosis or prognosis of HFpEF. Forest plot of NT-proBNP with estimated pooled sensitivity and specificity (A) and HSROC curve (B). (C) Random-effect model forest plot of hazard ratios for NT-proBNP to predict mortality, hospitalisation or composite event of both. CI, confidence interval; df, degree of freedom; HSROC, hierarchical summary receiver-operating characteristic.

In a subgroup analysis including only studies conducted in Europe (n=3),^s1, s2, s5^ the estimated pooled sensitivity was 0.68 (95% CI: 0.63 to 0.73) and specificity was 0.95 (95% CI: 0.93 to 0.98), similar to the primary analysis (Supplementary material online, *Figure S1A*). However, there was no heterogeneity in relation to the sensitivity (*I*^2^=0%), with substantial heterogeneity being reported in terms of the specificity (*I*^2^=60.8%) in this subgroup analysis. In another subgroup analysis including only patients from Asia (n=3), ^s3, s4, s6^ similar sensitivity (0.71, 95% CI: 0.65 to 0.76) and lower specificity (0.83, 95% CI: 0.76 to 0.89) were obtained (Supplementary material online, *Figure S1B*). However, heterogeneity was substantially high in terms of both sensitivity (*I*^2^=93.1%) and specificity (*I*^2^=75.6%).

Studies investigating NT-proBNP as a prognostic marker to predict the pre-specified primary outcomes in patients with HFpEF (n=8),^s19-s26^ were conducted in Asia (n=1),^s25^ Europe (n=3),^s19, s20, s26^ or other continents (n=4)^s21-s24^. Cut-off value for NT-proBNP varied across these eight studies (median: 763.77 pg/mL, range: 68.5 pg/mL-3606 pg/mL) as did the follow-up periods (median: 19.62 months, range: 6 months-5 years). Using data from all eight studies, the estimated pooled HRs were significant [mortality (n=7): 2.27, 95% CI: 1.69 to 3.06; hospitalisation (n=2): 3.54, 95% CI: 2.83 to 4.43; a composite event (n=8: 2.55, 95% CI: 2.13 to 3.05] (*Figure 2C*). Heterogeneity was prominent in studies assessing mortality (*I*^2^=87.6%) or composite event (*I*^2^=87.6%) with no heterogeneity being reported for hospitalisation (*I*^2^=0%). No evidence of systematic bias was present for mortality (Egger *P*=0.139) or composite event (*P*=0.182), however systematic bias was present for hospitalisation (hospitalisation: *P*<0.001).

### BNP as a biomarker of diagnosis and prognosis in HFpEF

The diagnostic accuracy of BNP in HFpEF (n=6),^s8-s13^ was assessed using a wide range of cut-off values (31 pg/mL to 353.6 pg/mL). Studies were conducted in Asia (n=3),^s8-s10^ North America (n=2) ^s11, s12^ and Europe (n=1) ^s13^. Both estimated sensitivity (0.81, 95% CI: 0.76 to 0.85) and specificity (0.86, 95% CI: 0.82 to 0.89) of BNP to diagnose HFpEF were highly reliable (*Figure 3A* and *3B*). Low heterogeneity was reported between these studies (sensitivity: *I*^2^=0%; specificity: *I*^2^=16.9%).

**Figure 3.**
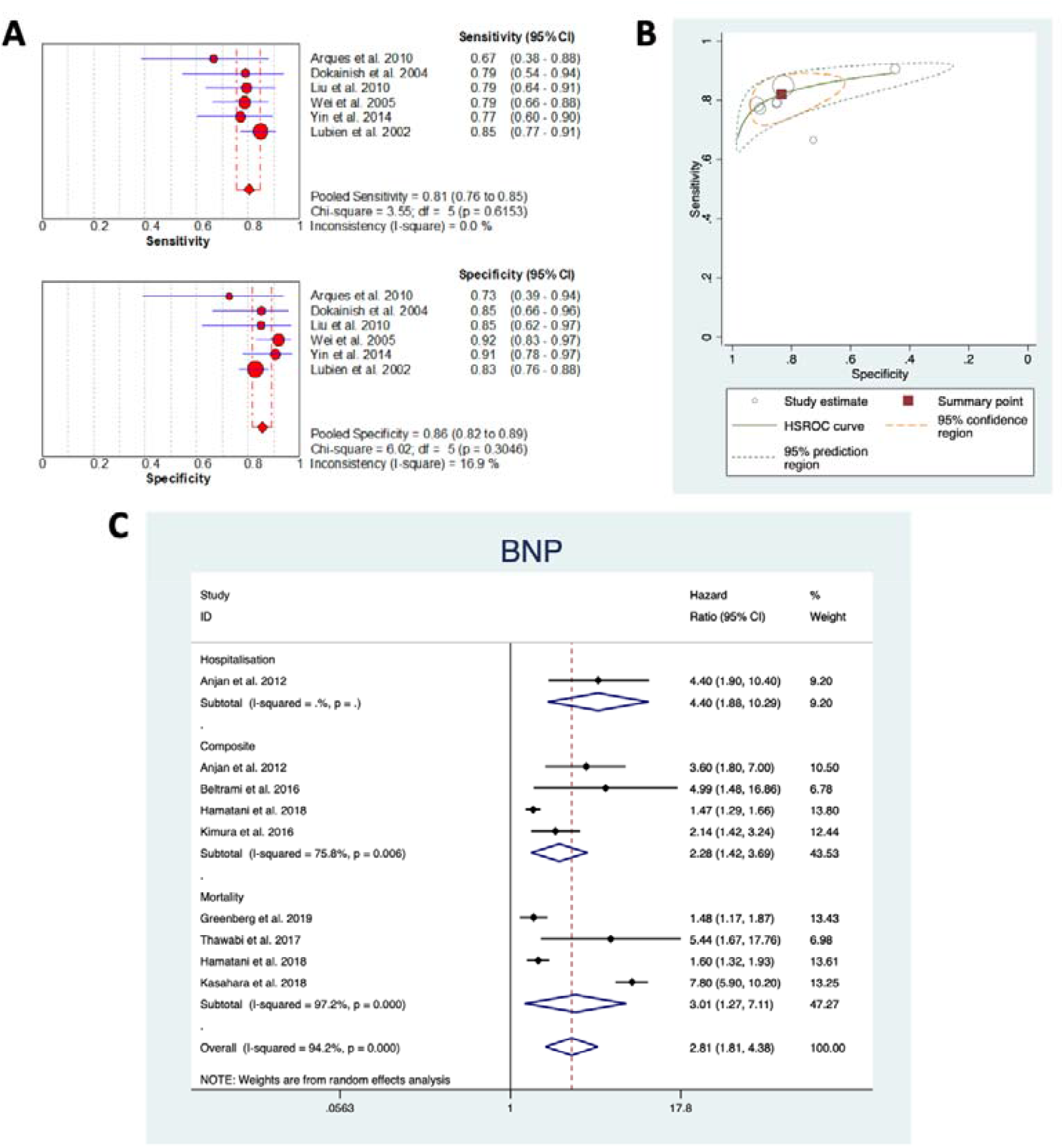
Sensitivity, specificity and hazard ratio of BNP extracted from all studies using BNP to diagnose HFpEF. Forest plot of BNP with estimated pooled sensitivity and specificity (A) and HSROC curve (B). (C) Random-effect model forest plot of hazard ratios for BNP to predict mortality, hospitalisation or composite event of both. CI, confidence interval; df, degree of freedom; HSROC, hierarchical summary receiver-operating characteristic.

In relation to the studies reporting the prognostic accuracy of BNP in patients with HFpEF (n=7),^s29-s35^ three ^s30, s31, s35^ were conducted in North America, three ^s32-s34^ in Asia, and one ^s29^ in Europe. Cut-off values of BNP used in these studies were varied (median: 287 pg/mL, range: 100 pg/mL to 1000 pg/mL) as did the follow-up periods (median: 21.83 months, range: 6 months-6.5 years). Only one study ^s30^ reported the accuracy of BNP to predict hospitalisation in patients with HFpEF. The estimated pooled HRs of BNP to predict mortality, or a composite event were 3.01 (95% CI: 1.27 to 7.11) and 2.28 (95% CI: 1.42 to 3.69), respectively (*Figure 3C*). Heterogeneity and bias were very high in relation to mortality (*I*^2^=97.2%, Egger *P*=0.607), whereas substantial heterogeneity and borderline bias were reported in predicting a composite event (*I*^2^= 75.8%, Egger *P*=0.071).

### Galectin-3 and sST2 as biomarkers of HFpEF diagnosis

All studies investigating Gal-3 as diagnostic biomarker (n=3),^s6-s8^ of HFpEF were conducted in Asia. Cut-off values ranged from 9.55 ng/mL to 20.12 ng/mL. Estimated pooled sensitivity and specificity of Gal-3 were 0.70 (95% CI: 0.63 to 0.75) and 0.78 (95% CI: 0.69 to 0.85) respectively (*Figure 4A*). Heterogeneity was large for both sensitivity (*I*^2^=86.7%) and specificity (*I*^2^=68.6%).

**Figure 4.**
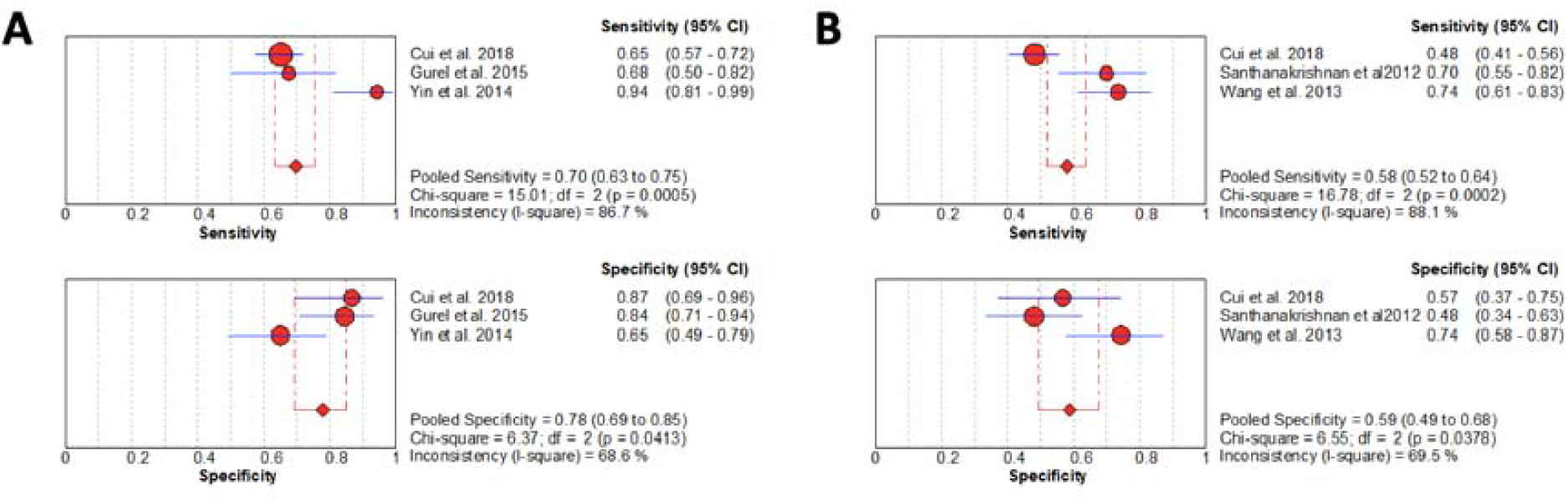
Diagnostic sensitivity and specificity of Gal-3 and sST2 extracted from all studies using Gal-3 to diagnose HFpEF. Forest plot of Gal-3 (A) and sST2 (B) with estimated pooled sensitivity and specificity. CI, confidence interval; df, degree of freedom.

Studies investigating sST2 to diagnose HFpEF (study n=3),^s3, s6, s14^ were all also conducted in Asia. Cut-off values were highly varied across the studies and ranged from 68.6 pg/mL to 26.47 ng/mL. Limited diagnostic sensitivity (0.58, 95% CI: 0.52 to 0.64) and specificity (0.59, 95% CI: 0.49 to 0.68) of sST2 to diagnose HFpEF were observed (*Figure 4B*). Heterogeneity was also significant in terms of both sensitivity (*I*^2^=88.1%) and specificity (*I*^2^=69.5%).

### Emerging biomarkers of HFpEF diagnosis and prognosis

Other diagnostic and prognostic biomarkers were also appraised in this study, however, meta-analysis was not conducted on these markers due to the granularity of data (n<3). As such, we presented the original sensitivity and specificity from each paper in relation to the following diagnostic biomarkers: tissue inhibitor matrix metalloproteinase 1 (TIMP1), adiponectin, angiogenin, soluble glycoprotein 130 (sgp130), heat shock protein 27 (hsp27), cardiac bridging integrator 1 (cBIN1), troponin-T (TnT), and growth differentiator factor 15 (GDF15); and also directly extracted HRs of the following prognostic biomarkers: midregional pro-A-type natriuretic peptide (MR-proANP), midregional proadrenomedullin (MR-proADM), Gal-3, sST2, cystatin C (CysC), tumour necrosis factor α (TNF-α), carbohydrate antigen-125 (CA-125), cBIN1, parathyroid hormone (PTH), troponin-I (TnI), TnT, C-reactive protein (CRP), thrombospondin-2 (TSP-2), copeptin, and migration inhibitory factor (MIF) (Supplementary material online, *Figure S2A* and *B*). In terms of diagnosis, adiponectin appears to have the highest sensitivity and specificity,^S18^ with varied prognostic potential compared to the rest of the biomarkers.

## DISCUSSION

This meta-analysis is the first systematic synthesis of available clinical observational studies evaluating diagnostic and prognostic accuracy of circulating biomarkers in HFpEF. The main findings of this study are: (i) NPs are the most reported biomarkers for diagnosis and prognosis of HFpEF, (ii) NT-proBNP is the biomarker with the highest diagnostic specificity, whereas BNP is the most reliable in diagnosing HFpEF in terms of both sensitivity and specificity, and (iii) NPs could also be used for predicting mortality, hospitalisation or a composite event of both events in HFpEF patients, however the heterogeneity was very high between the studies.

Although consensus guidelines have acknowledged the incremental value of NPs in diagnosis ^5,6^ and prognosis ^6^ of HFpEF, the diagnostic and prognostic accuracy of NPs in HFpEF have never been systematically reviewed in the context of both diagnosis and prognosis. We found that NT-proBNP has the highest specificity but modest sensitivity to diagnose HFpEF. This modest sensitivity was also confirmed by a retrospective study in which low (below cut-off) NT-proBNP levels were reported in about a half of the HFpEF cohort.^11^ According to our meta-analysis results, BNP may be the most reliable biomarker in HFpEF diagnosis due to its satisfactory sensitivity and specificity, as well as the reliability of its diagnostic potential over a large spectrum of cut-off values.

With respect to the predictive potential of NPs in HFpEF prognosis, both NT-proBNP and BNP were associated with increased risk of mortality, hospitalisation or a composite event, by over 2-fold. However, we are unable to determine which one is superior due to inconsistency in the results. In terms of NT-proBNP, three studies ^s22, s25, s26^ showed substantial variability in the results, demonstrated by substantial heterogeneity. There were inconsistencies in either patient characteristics, LVEF, follow-up periods or cut-off values in these three studies (Table 2). For BNP, the heterogeneity appears to be dependent on the study design resulting in variable mortality rates of HFpEF. Mortality rates of HFpEF patients in RCTs has been found to be significantly lower than that of non-RCTs, which may affect the HRs.

**Table 1.**
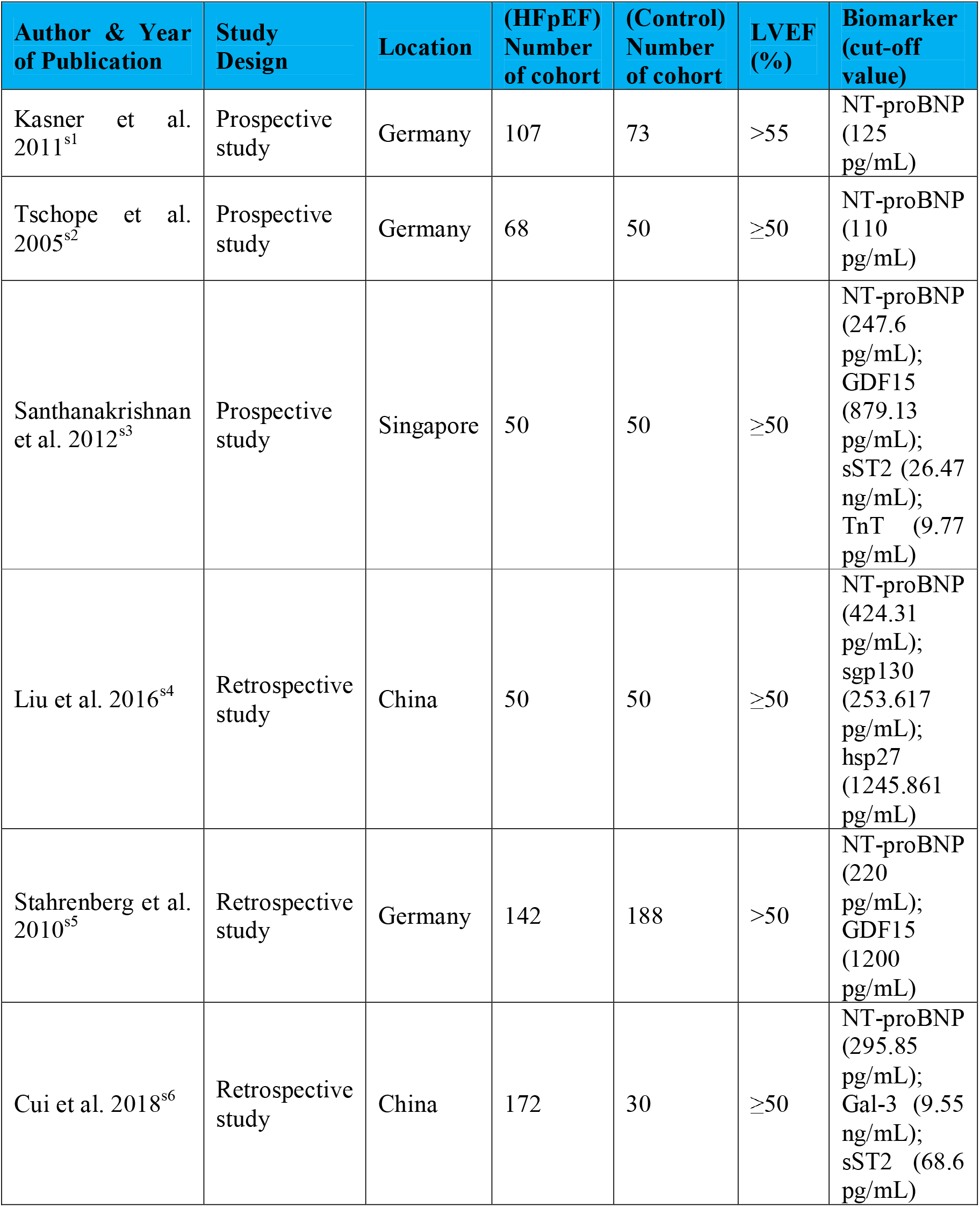
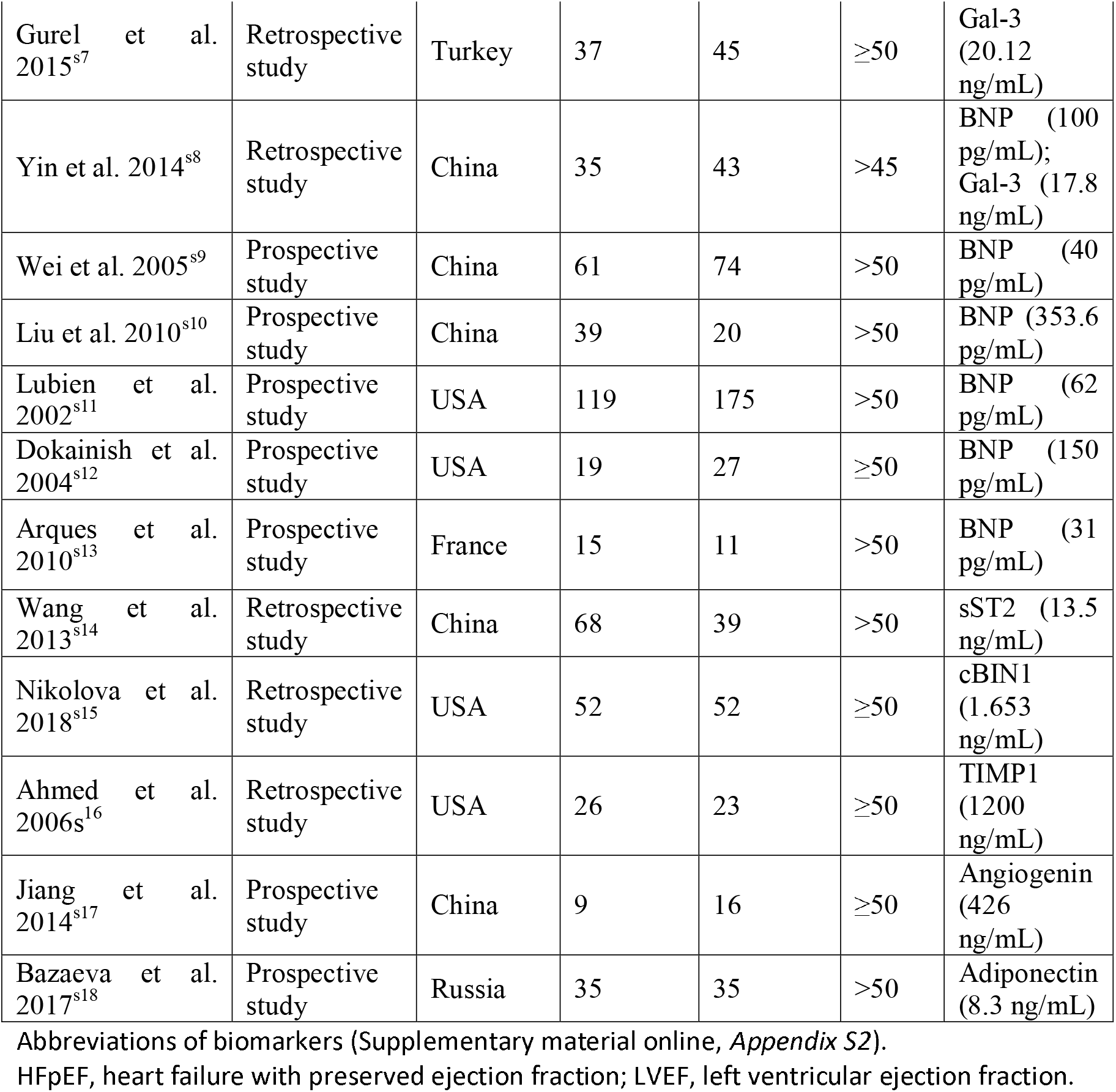
Study characteristics of included studies in relation to HFpEF diagnostic biomarkers

| Author & Year of Publication | Study Design | Location | (HFpEF) Number of cohort | (Control) Number of cohort | LVEF (%) | Biomarker (cut-off value) |
| --- | --- | --- | --- | --- | --- | --- |
| Kasner et al. 2011<sup>s1</sup> | Prospective study | Germany | 107 | 73 | >55 | NT-proBNP (125 pg/mL) |
| Tschope et al. 2005<sup>s2</sup> | Prospective study | Germany | 68 | 50 | ≥50 | NT-proBNP (110 pg/mL) |
| Santhanakrishnan et al. 2012<sup>s3</sup> | Prospective study | Singapore | 50 | 50 | ≥50 | NT-proBNP (247.6 pg/mL); GDF15 (879.13 pg/mL); sST2 (26.47 ng/mL); TnT (9.77 pg/mL) |
| Liu et al. 2016<sup>s4</sup> | Retrospective study | China | 50 | 50 | ≥50 | NT-proBNP (424.31 pg/mL); sgp130 (253.617 pg/mL); hsp27 (1245.861 pg/mL) |
| Stahrenberg et al. 2010<sup>s5</sup> | Retrospective study | Germany | 142 | 188 | >50 | NT-proBNP (220 pg/mL); GDF15 (1200 pg/mL) |
| Cui et al. 2018<sup>s6</sup> | Retrospective study | China | 172 | 30 | ≥50 | NT-proBNP (295.85 pg/mL); Gal-3 (9.55 ng/mL); sST2 (68.6 pg/mL) |
| Gurel et al. 2015<sup>s7</sup> | Retrospective study | Turkey | 37 | 45 | ≥50 | Gal-3 (20.12 ng/mL) |
| Yin et al. 2014<sup>s8</sup> | Retrospective study | China | 35 | 43 | >45 | BNP (100 pg/mL); Gal-3 (17.8 ng/mL) |
| Wei et al. 2005<sup>s9</sup> | Prospective study | China | 61 | 74 | >50 | BNP (40 pg/mL) |
| Liu et al. 2010<sup>s10</sup> | Prospective study | China | 39 | 20 | >50 | BNP (353.6 pg/mL) |
| Lubien et al. 2002<sup>s11</sup> | Prospective study | USA | 119 | 175 | >50 | BNP (62 pg/mL) |
| Dokainish et al. 2004<sup>s12</sup> | Prospective study | USA | 19 | 27 | ≥50 | BNP (150 pg/mL) |
| Arques et al. 2010<sup>s13</sup> | Prospective study | France | 15 | 11 | >50 | BNP (31 pg/mL) |
| Wang et al. 2013<sup>s14</sup> | Retrospective study | China | 68 | 39 | >50 | sST2 (13.5 ng/mL) |
| Nikolova et al. 2018<sup>s15</sup> | Retrospective study | USA | 52 | 52 | ≥50 | cBIN1 (1.653 ng/mL) |
| Ahmed et al. 2006<sup>s16</sup> | Retrospective study | USA | 26 | 23 | ≥50 | TIMP1 (1200 ng/mL) |
| Jiang et al. 2014<sup>s17</sup> | Prospective study | China | 9 | 16 | ≥50 | Angiogenin (426 ng/mL) |
| Bazaeva et al. 2017<sup>s18</sup> | Prospective study | Russia | 35 | 35 | >50 | Adiponectin (8.3 ng/mL) |

Abbreviations of biomarkers (Supplementary material online, *Appendix S2*).
HFpEF, heart failure with preserved ejection fraction; LVEF, left ventricular ejection fraction.

**Table 2.**
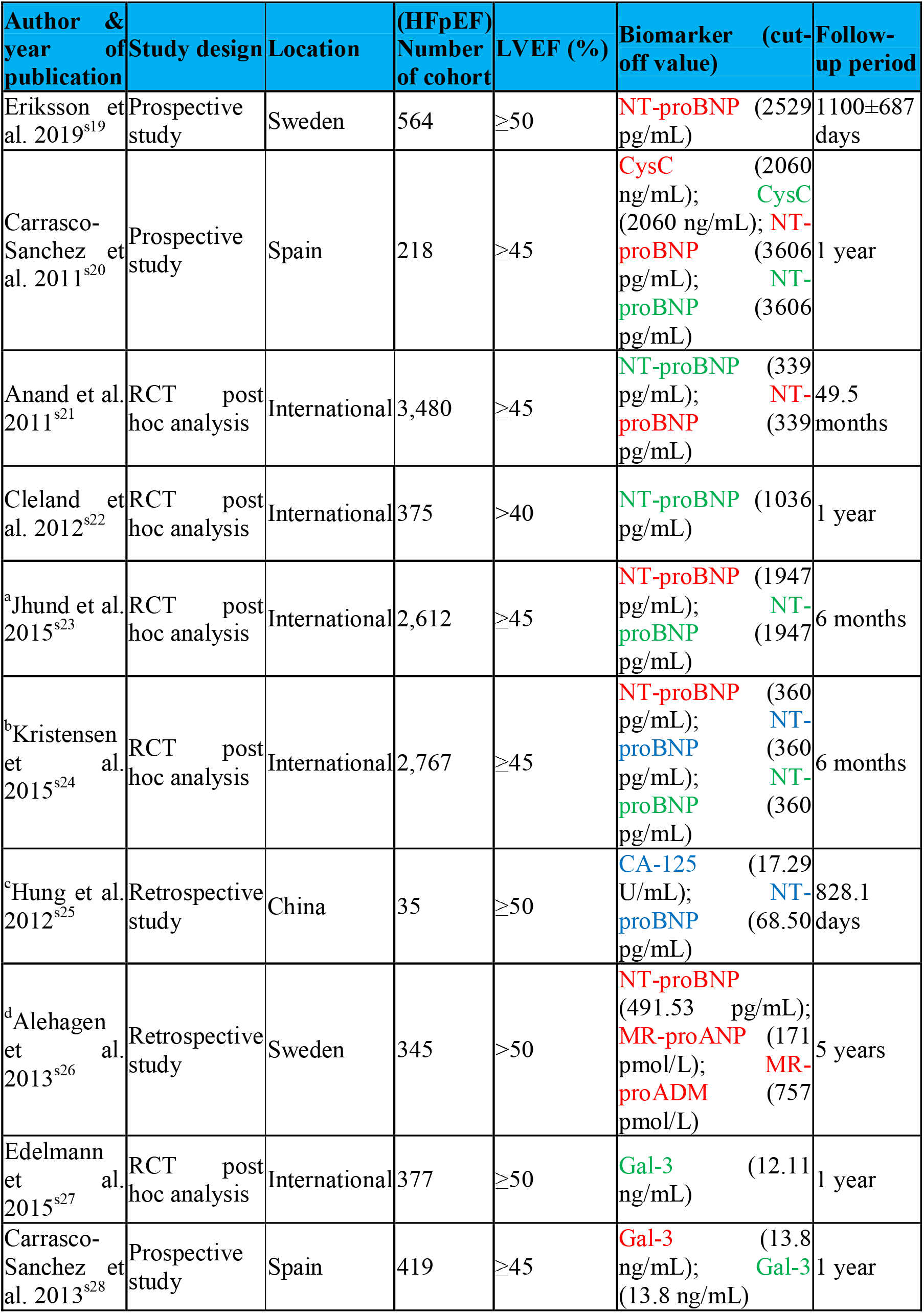
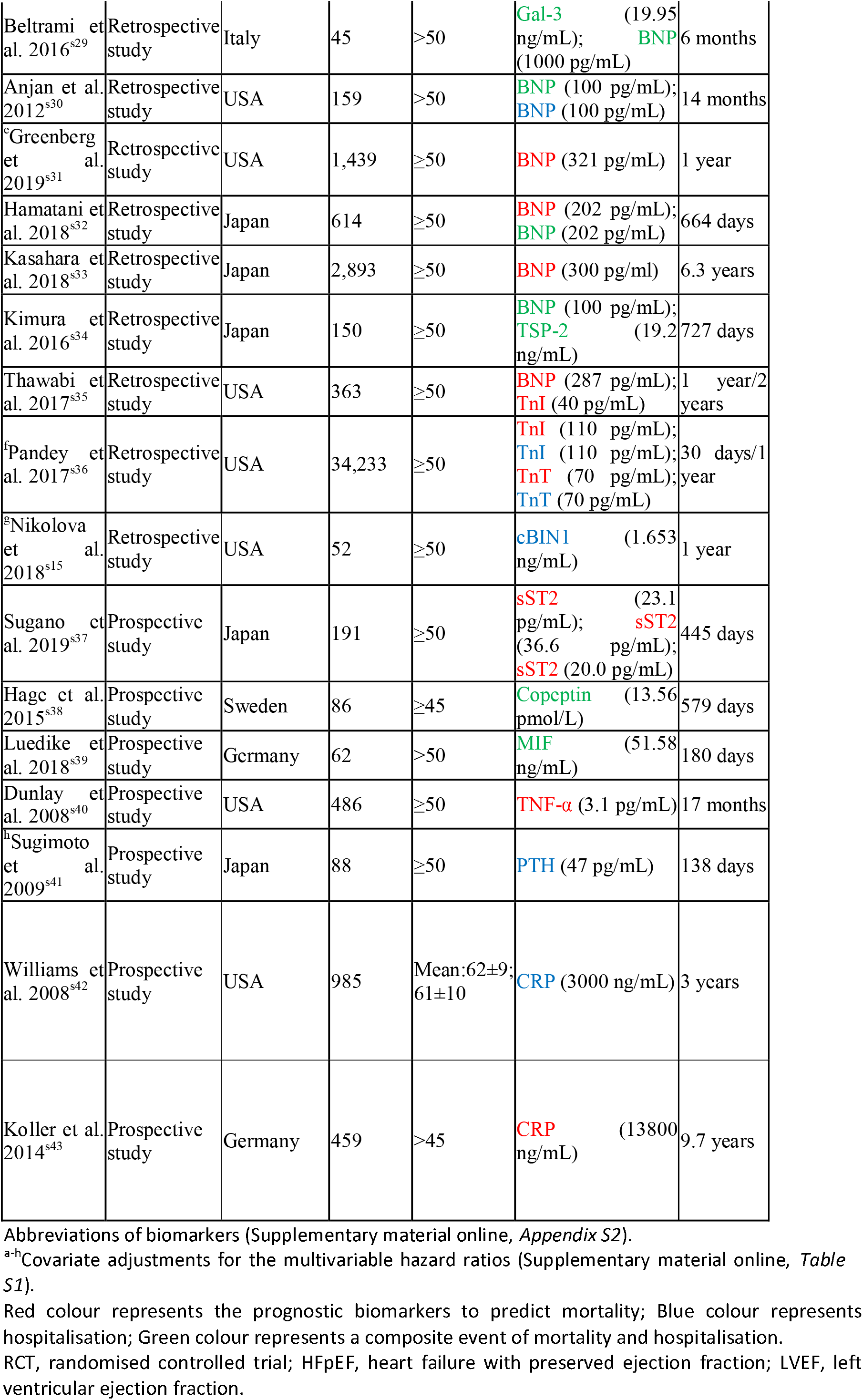
Study characteristics of included studies in relation to HFpEF prognostic biomarkers

In a previous study comparing biomarkers in HFpEF and HFrEF using a novel heatmap method, NPs demonstrated a strong association with HFrEF, whereas biomarkers of inflammation and angiogenesis were predominantly associated with HFpEF.^12^ The latter was also confirmed in a recent study. ^13^ Gal-3, a well-characterised inflammatory marker, has shown potential as a promising biomarker of HFpEF in pre-clinical models ^14-15^ and according to the epidemiological evidences ^16, 17^. Nevertheless, in this meta-analysis a limited diagnostic value of Gal-3 was demonstrated in HFpEF, based on observational clinical studies. High heterogeneity was present between studies investigating the diagnostic potential of Gal-3, which could have been caused by the wider range of LVEF (>45%) used to recruit patients in one included study,^s8^ that may have resulted in the inclusion of patients with HF with mid-range ejection fraction as part of this small HFpEF cohort (n=35). In addition, unlike NPs, which show differentially elevated concentrations in HFpEF and HFrEF,^18-20^ Gal-3 concentrations are unable to distinguish HFpEF from HFrEF,^19^ suggesting lower diagnostic value of Gal-3 than that of NPs. Despite its limited diagnostic value, Gal-3 could represent a promising prognostic marker in HFpEF. Although we did not find a sufficient number of studies to conduct a meta-analysis in relation to Gal-3’s prognostic potential using our inclusion criteria, a number of other studies have reported potent prognostic value of Gal-3 in HFpEF.^21-25^ However, more studies are needed to validate this hypothesis, given the fact that Gal-3 level is a confounder of NT-proBNP and renal function.^25, 26^

Similarly, although promising results have been reported with sST2 in HFpEF,^27, 28^ in our meta-analysis limited diagnostic and prognostic values of sST2 have been demonstrated. Our results revealed that the diagnostic accuracy of sST2 in HFpEF is highly dependent on its cut-off values, hence heterogeneity was substantial when we pooled sensitivities and specificities from different studies. In addition, sST2 is unable to distinguish HFpEF cohorts from healthy individuals after adjusting for age, sex and other clinical covariates,^29^ which may be caused by the lack of association of sST2 with LV function and structure.^30^ We suggest that the utility of NPs appears currently superior to Gal-3 and sST2, in terms of HFpEF diagnosis.

### Study limitations

Due to the granularity of data, we were unable to assess the diagnostic and prognostic accuracy of other emerging biomarkers, which should be investigated in the future through meta-analysis when there are sufficient number of studies investigating these biomarkers in diagnosis and prognosis of HFpEF. We also acknowledged that there is high heterogeneity in most of the meta-analyses performed except for the diagnostic potential of BNP in HFpEF, which should be addressed in future meta-analyses incorporating further and more homogeneous studies. This is likely caused by the variability in study designs, biomarker cut-off values and follow-up periods.

## CONCLUSION

HFpEF comprises approximately half of all patients with HF, with similar morbidity and mortality incidence, yet it is poorly understood and pharmacologically managed. Due to the lack of understanding of the pathogenesis of HFpEF, and diagnostic challenges, a delay in diagnosis and treatment is common, which can lead to worse outcomes for patients with HFpEF. Accurate biomarkers are clinically important to improve both diagnosis and prognosis of HFpEF, emphasising urgent need for biomarker discovery and validation. Nevertheless, the status of biomarkers for diagnosis and prognosis of HFpEF, demonstrated in this meta-analysis, suggests that NPs, particularly BNP, remain the most reliable biomarkers in diagnosing HFpEF, with some potential for HFpEF prognosis.

## Data Availability

All the data is included in the paper.

## FUNDING

This project was funded through the Honour’s student fund provided by the Faculty of Science, University of Technology Sydney.

## CONFLICT OF INTEREST

Authors declare no conflict of interest.

## Notes

### Competing Interest Statement

The authors have declared no competing interest.

